# Inspiratory Strength Training in Pediatric Cardiac Critical Care: A Retrospective Cohort Study

**DOI:** 10.64898/2026.08.13.26360419

**Authors:** Jessica Braun Cornman, A. Daniel Martin, Juliette Clavier, Joseph Philip, Giles J Peek, Jeffrey P. Jacobs, Mark S. Bleiweis, Barbara K. Smith

## Abstract

**Background:** Prolonged mechanical ventilation (MV) is associated with inspiratory muscle weakness and difficulty weaning from respiratory support. While inspiratory strength training (IST) has demonstrated efficacy in adult critical care populations, the data in pediatric cardiac critical care remains limited. We sought to evaluate the feasibility, safety, and physiologic response to IST in children in the pediatric cardiac intensive care unit (PCICU).

**Methods and Results:** We performed a single-center retrospective cohort study of children with congenital heart disease referred for IST between January 2015 and August 2021. Feasibility was defined as completion of ≥1 IST session following referral. Safety outcomes included physiologic events documented during IST sessions. Changes in maximal inspiratory pressure (MIP) were assessed in patients who completed ≥2 IST sessions. Of 105 eligible patients, 93 (89%) successfully completed at least 1 IST session. Across 389 IST sessions, monitoring events included pre-oxygenation (62%), desaturations (13%), bradycardia (7%), and hypertension (2%). All events were transient and did not require escalation of care. 84% of patients were successfully liberated from MV after a median of 2 (IQR 1-4) IST sessions. Among patients completing ≥2 sessions, MIP improved significantly over time (p < 0.0001). Improvements were observed in both patients who did and did not wean from MV. Patients who failed to wean had longer ventilator exposure prior to IST initiation and greater sedation at baseline.

**Conclusions:** IST was feasible and well tolerated in this medically complex PCICU cohort. High completion rates and improvements in MIP support IST as a clinically deliverable intervention that can improve inspiratory muscle strength during critical illness.

## Introduction

Congenital heart defects (CHDs) are the most common type of congenital anomaly and exhibit wide variability in clinical presentation. Many infants and children with CHDs will require surgical repair or palliation, accompanied by mechanical ventilation (MV). The requirement for MV is variable and dependent on hemodynamic stability, disease severity, and weaning tolerance. Although lifesaving, prolonged mechanical ventilation is associated with a rapid reduction in diaphragm force production and cross-sectional area, known as ventilator-induced diaphragm dysfunction. This dysfunction has been strongly associated with difficulty weaning from mechanical ventilation and delayed extubation. Respiratory muscle weakness may persist beyond the intensive care unit (ICU) stay and is correlated with long-term morbidity. Respiratory muscle weakness has been reported to persist into late childhood and early adulthood in children with Fontan physiology. Prolonged respiratory muscle weakness beyond ICU discharge underscores the need for targeted interventions during critical illness that can improve motor recovery.

Inspiratory strength training (IST) has demonstrated strong clinical efficacy in adult ICU populations, including improvements in maximal inspiratory pressure, reduced time to ventilator liberation, improved functional aerobic capacity, and reduced postoperative pulmonary complications, particularly in cardiac surgical patients.In pediatric populations, inspiratory strength training has been shown to be safe and effective in improving maximal inspiratory pressure across a range of diagnoses; however, these studies are limited to very specific populations, and the majority have taken place outside of the ICU.Evidence describing inspiratory strength training in pediatric critical care remains sparse and largely limited to case reports and small case series.

Based on strong adult safety and efficacy data and our early pediatric experience, our institution implemented inspiratory strength training (IST) as a standard physical therapy intervention in our Pediatric ICU (PICU) and Pediatric Cardiac ICU (PCICU) for children with new-onset ventilator dependence and clinical concerns for weaning difficulties. In this retrospective cohort study, we aimed to describe IST utilization patterns; evaluate feasibility (delivery of at least 1 session following referral); evaluate safety (frequency and severity of physiologic events during IST sessions); and characterize changes in maximal inspiratory pressure (MIP) from baseline to final measurement among patients completing 2 or more sessions. As an exploratory analysis, we compared clinical characteristics between patients who were and were not liberated from invasive mechanical ventilation (off invasive MV for 72 hours) during the hospitalization. We hypothesized that IST is feasible, well tolerated, and associated with improvements in MIP over the course of training.

## Methods

### Study design and setting

We conducted a single-center retrospective cohort study of children in the pediatric critical care units of a tertiary care center. Admissions were identified using the institutional data repository to query electronic medical records for patients with an order for “PT eval and treat IMT” who were admitted to our pediatric intensive care unit (PICU) or our pediatric cardiac intensive care unit (PCICU) between January 1, 2014, and August 31, 2021. IST was implemented as a standard clinical practice in 2014 for patients with new-onset ventilator dependence and perceived difficulty with weaning. When the program was initially established, IST was performed by a single trained physical therapy clinician-scientist. Starting in 2016, two additional pediatric physical therapists were trained to perform IST evaluation and intervention. This core group of specially trained therapists provides routine weekday and weekend coverage for IST. The University of Florida Institutional Review Board approved the research procedures (IRB202101627). Because the study methods involved a retrospective review of standard practice, informed consent from a parent or caregiver was not required.

### Subjects

Admissions were eligible for inclusion and data extraction if the patient was admitted to the PCICU or PICU services between January 1, 2014, and August 31, 2021, were under the age of 18 at the time of the evaluation, and required positive pressure ventilatory support. Admissions were excluded if the patient had a cervical spinal cord injury, required full-time mechanical ventilation at baseline, or underwent multiple distinct courses of IST during the same hospitalization (e.g., following both ventricular assist device placement and heart transplantation). Approximately 226 individual admissions received a referral for IST in both the PICU and PCICU. The current study focused on only those with a diagnosis that was cardiac in nature; 105 met criteria for this cardiac subanalysis. Both invasive and noninvasive ventilation-dependent subjects were included, with 102 subjects invasively ventilated at evaluation (97%). For included admissions, baseline MIP was obtained from the first recorded session, and final MIP taken at the last recorded session prior to extubation or IST discontinuation.

### Maximal Inspiratory Pressure Measurement

MIP was measured by placing a unidirectional valve placed in series with the endotracheal or tracheostomy tube to transiently occlude inspiration and maximally stimulate inspiratory effort. For individuals on noninvasive support, an oronasal mask was utilized as the interface for the unidirectional valve. To minimize physiologic stress, each session included 3-4 occlusion trials, each limited to 15-20 seconds depending on patient tolerance. Between each occlusion trial, subjects received approximately 2 minutes of rest. The highest value obtained across all trials was recorded for MIP analysis.

### Inspiratory Strength Training

While various respiratory training devices are commercially available, they are unsuitable for many pediatric patients. Sedation or pain medications blunt the inspiratory drive and further restrict older children’s ability to follow training cues. Moreover, the large dead space of commercial devices outweighs the small respiratory volumes generated by infants and small children. Most IST training sessions in the PCICU were performed using an occlusive training technique, as described in a case report by Smith et al..^20^ A small subset of patients used a threshold IST device to train against submaximal inspiratory training loads.

### Data retrieval

Two investigators were trained in data extraction, reviewed each electronic medical record, and recorded data using a standardized data collection spreadsheet. Extracted variables included demographics, congenital heart defect as documented by cardiology, illness severity (PRISM V), ventilator characteristics, sedation level at IST initiation (State Behavioral Scale), IST session details and MIP values, and clinical outcomes. Quality review of a subset of records revealed an inter-rater agreement of 93%. De-identified data from each subject were compiled into a summary spreadsheet for final data analysis.

### Data analysis

Demographics and clinical characteristics were summarized using descriptive statistics (mean [SD], median [IQR], and frequency[%]). Feasibility was assessed by completion of IST evaluation and delivery of one IST session following referral. Due to the retrospective nature of the study and the use of clinical medical records, we were unable to track missed or deferred treatment sessions. The primary safety outcome was the frequency and type of physiologic events documented during IST sessions and whether any intervention beyond standard bedside care was warranted. Liberation from invasive mechanical ventilation was defined as being off invasive mechanical ventilation for 72 hrs. Initial physiologic response was defined as the change in maximal inspiratory pressure from baseline to the final recorded value among patients completing 2 IST sessions, assessed using a mixed-effects analysis. Unpaired t-test and Mann-Whitney U tests were used to assess group differences between those who successfully weaned from mechanical ventilation and those who did not, as well as between those who successfully performed IST and those who did not. Significance was based on a two-sided α level of .05. Statistical analyses were performed using GraphPad Prism version 10.6.1.

## Results

### Demographic

From January 2014 to August 2021, 226 patients were referred from a designated health record physical therapy consult for IST. Of these, 105 met inclusion criteria for this study. Of the 105, 93 (89%) received at least 1 session of IST, and 12 (11%) did not because extubation occurred prior to physical therapy evaluation (Figure 1). Among all admissions, 88 patients were liberated from invasive mechanical ventilation, and 17 were not (Table 1). In the weaned group, 28 (32%) had acyanotic defects, 52 (59%) had cyanotic or critical defects, and 8 (9%) were under evaluation for or recieved a VAD or heart transplant. In the unweaned group, 7 (41%) had acyanotic defects, 8 (47%) had cyanotic or critical defects, and 2 (12%) under evaluation for or recieved a VAD or heart transplant. There were no significant differences in patient weight or age between the weaned and unweaned groups at the outset of IST (weight: p=0.12, age: p=0.56). Hospital courses were largely similar between the weaned and unweaned groups, except for days of mechanical ventilation prior to IST, with the weaning group requiring far fewer days of mechanical ventilation prior to referral for IST (p<0.0001), and for the state behavioral scale (SBS) score, with the unweaned group reported as more sedated at initiation of IST (p=0.023) (Table 2). Survival to hospital discharge was significantly greater in the weaned subjects (88.6%) compared to the unweaned subjects (41.2%) (Fisher’s exact test, p<0.0001), with the majority of deaths attributed to complex cardiac disease. The odds of survival were 11.1-fold higher in weaned subjects (OR 11.14, 95% CI 3.35-36.10). Among the 7 surviving unweaned subjects, weaning failure was attributed to tracheobronchomalacia (n=4), upper airway dysfunction (n=2), and severe bronchopulmonary dysplasia (n=1).

**Figure 1.**
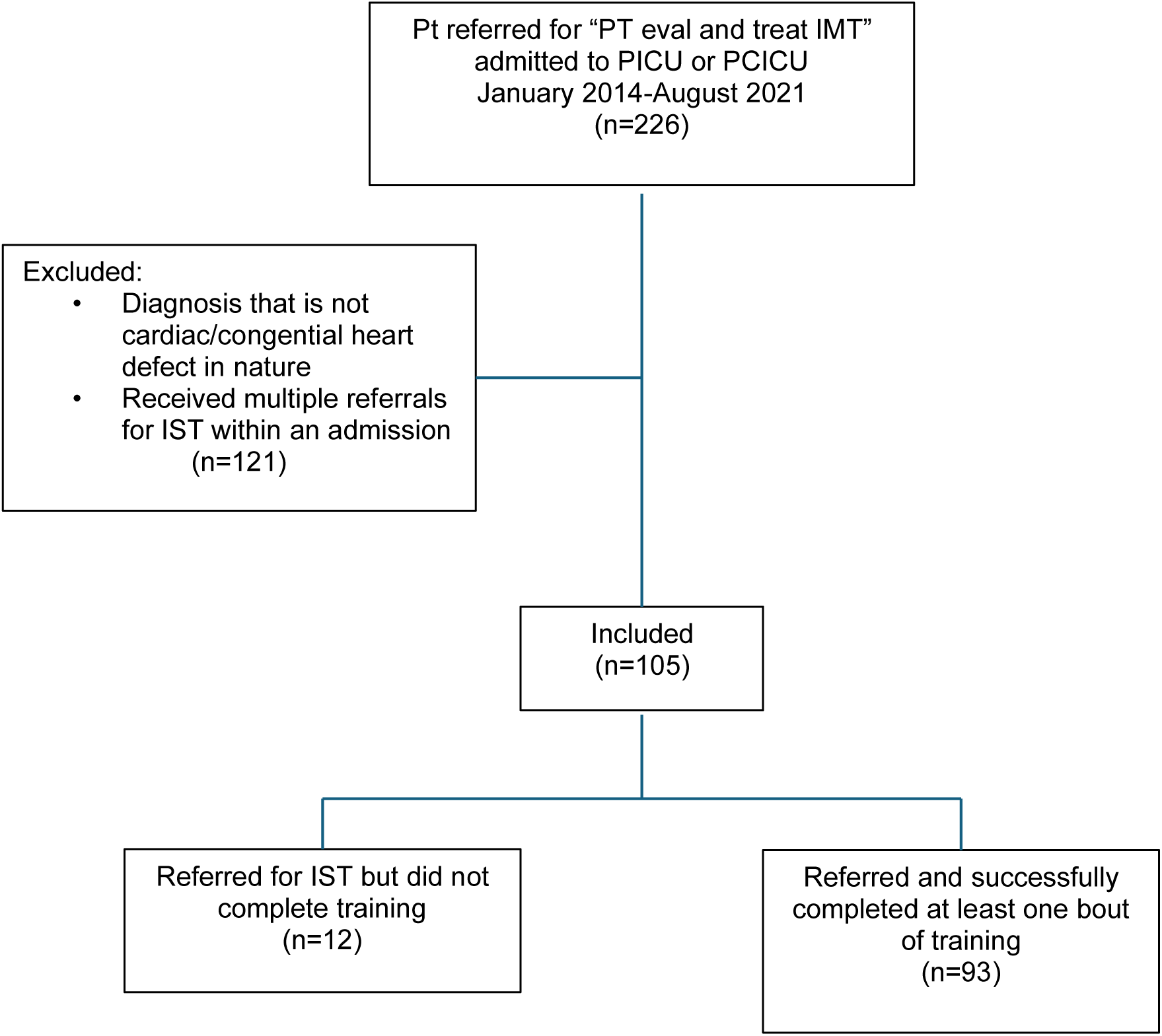
Study population selection flow diagram. Flow diagram depicting patient screening, eligbility assessment, and final inclusion. Reasons for exclusion at each stage are shown with the final cohort stratified into weaned and unweaned group.

**Table 1.** Demographic and Clinical Characteristics Stratified by Weaning Status. . Continuous variables are presented as mean ± SD, categorical variables are presented as counts. Comparisons between groups were performed using unpaired t-tests or Fishr’s exact tets, as appropriate. PRISM indicates Pediatric Risk of Mortality.

| Demographics | Weaned (n=88) | Unweaned (n=17) | Significance |
| --- | --- | --- | --- |
| **Age (months) (mean)** | 34.53 ± 64.51 | 4.118 ± 7.305 | p=0.5647 |
| **Weight (kg)** | 18.78 ± 33.07 | 6.084 ± 9.689 | p=0.2579 |
| **PRISM IV Score** | 19.54 ± 20.81 | 22.53 ± 23.74 | p=0.5700 |
| **Race/Ethnicity** |  |  | p=0.4346 |
| **White** | 49 | 9 |  |
| **Black** | 28 | 5 |  |
| **Hispanic** | 5 | 0 |  |
| **Asian** | 2 | 1 |  |
| **Other** | 4 | 2 |  |
| **Cardiac Defect** |  |  | p=0.6563 |
| **Cyanotic/Critical** | 52 | 7 |  |
| **Acyanotic** | 28 | 8 |  |
| **VAD/Transplant** | 8 | 2 |  |

**Table 2.** Hospitalization Characteristics of Weaned and Unweaned Patients. Values are presented as mean±SD. P values reflect between -group comparisons. MV indicated mechanical ventilation; IST, inspiratory strength training; SBS, State Behavioral Scale; MIP, maximal inspiratory pressure; SpO_2_/FiO_2_, oxygen saturation to fraction of inspired oxygen ratio.

| Hospitalization Characteristics | Weaned (n=88) | Unweaned (n=17) | Significance |
| --- | --- | --- | --- |
| **Hospital Days Prior to IST** | 41.39 ± 52.15 | 50.65 ± 42.43 | p=0.0727 |
| **MV days Prior to IST** | 19.05 ± 20.98 | 54.41 ± 44.13 | p<0.0001* |
| **SBS at IST Initiation** | -0.1923 ± 0.8836 | -0.7647 ± 1.091 | p=0.0214* |
| **Procedures during IST** | 5.931 ± 10.29 | 4.647 ± 9.493 | p=0.0864 |
| **Hospital Length of Stay** | 103.3 ± 80 | 133.5 ± 92.13 | p=0.2263 |
| **Baseline MIP** | 54.02 ± 22.71 | 50.04 ± 22.08 | p=0.5139 |
| **SpO<sub>2</sub>/FiO<sub>2</sub> Ratio at IST** | 284.1 ± 101.1 | 302.4 ± 91.94 | p=0.4946 |

### Feasibility

Of the 105 patients referred from IST during the study period, 93 (89%) successfully completed at least one session of IST. Every patient who did not receive IST was successfully extubated prior to the physical therapy evaluation, and approximately half of the missed evaluations occurred when referrals were placed on the weekend.

### Safety

The 93 individual patients who successfully completed IST underwent a cumulative total of 389 IST sessions. Monitoring events were documented for each IST session. These events included a preemptive oxygen boost on the ventilator that continued when the patient was placed back on the ventilator, suctioning before or during a session, desaturation, bradycardia, and hypertension. The majority of monitoring events were FiO2 boosts from the ventilator (62% of reported events), followed by need for suctioning (16%) and transient desaturation (13%). Bradycardia (7%) and hypertension (2%) were infrequent and transient (Figure 2). All reported monitoring events self-resolved, and none required intervention by physicians or advanced practice providers or an escalation from the current level of care.

**Figure 2.**
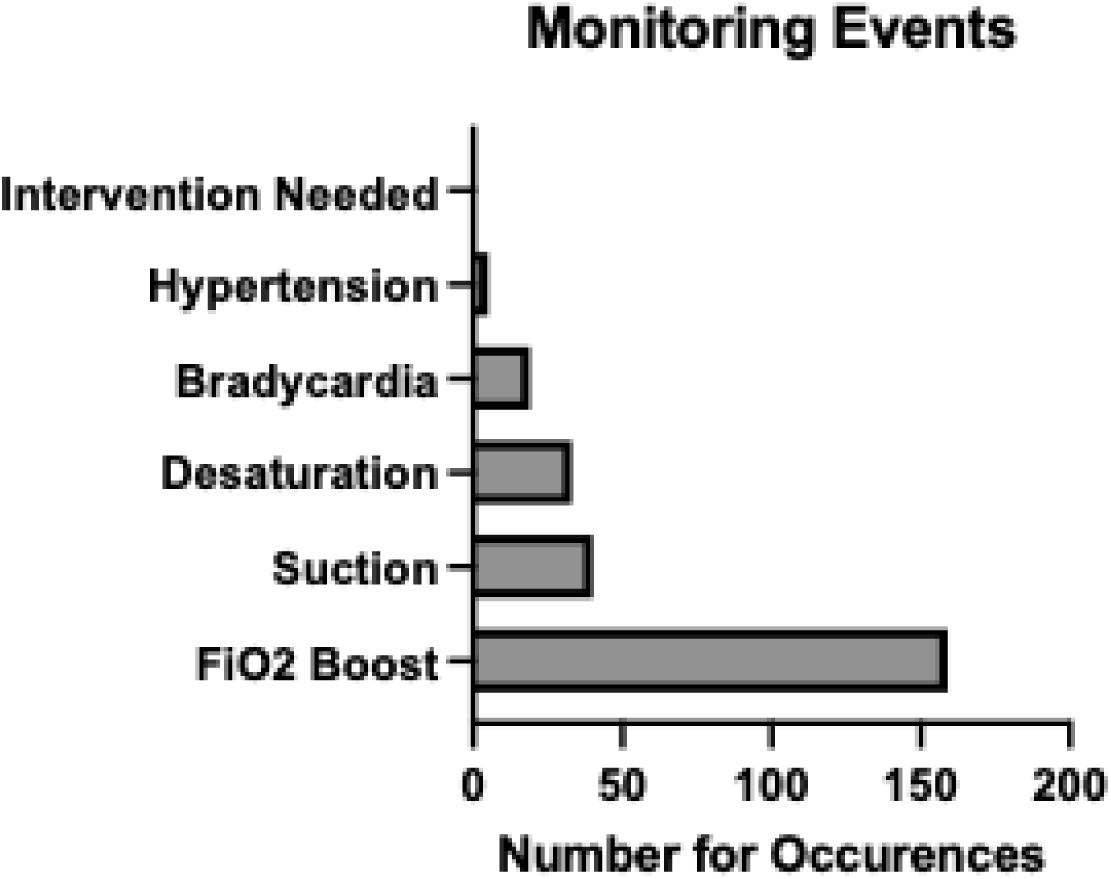
Monitoring Events During Inspiratory Strength Training Sessions. Graphical representation of the distribution and frequency of monitoring events observed during inspiratory strength training sessions. Bars represent the number of recrded events across the study period.

### Initial Physiologic Response

Patients who were liberated from invasive mechanical ventilation received significantly fewer IST sessions (p=0.003) than children who did not wean; a median of 2 sessions was required to achieve ventilator weaning (IQR 1-4) (Figure 3). Patients who failed to wean from mechanical ventilation had significantly more ventilator days (median 46 days [IQR 22.5-77.5; n=17]) prior to initiation of IST than those who weaned (median 11 days [IQR 5-25; n=87]) (Mann-Whitney U=273, p<0.0001). Despite prolonged mechanical ventilation in both the weaned and unweaned groups, a mixed-effects model demonstrated a significant main effect of time on MIP (F(1,60)=17.79, p<0.0001) for individuals receiving 2 or more IMT sessions. There was no significant effect of weaning status (F(1,61)=0.38, p=0.54) and no significant time by weaning interaction (F(1,60)=0.16, p=0.69), indicating MIP improved significantly for patients receiving 2 or more IST sessions regardless of weaning status. Those who weaned improved their MIP by an average of 15.24 cmH2O (SD 20.39), while the group that failed to wean improved their MIP by 12.32 cmH2O (SD 21.6). MIP also improved significantly over the course of training in patients receiving two or more sessions, regardless of survival status (p<0.0001). Despite various complexities and outcomes, MIP improved in all groups (Figure 4).

**Figure 3.**
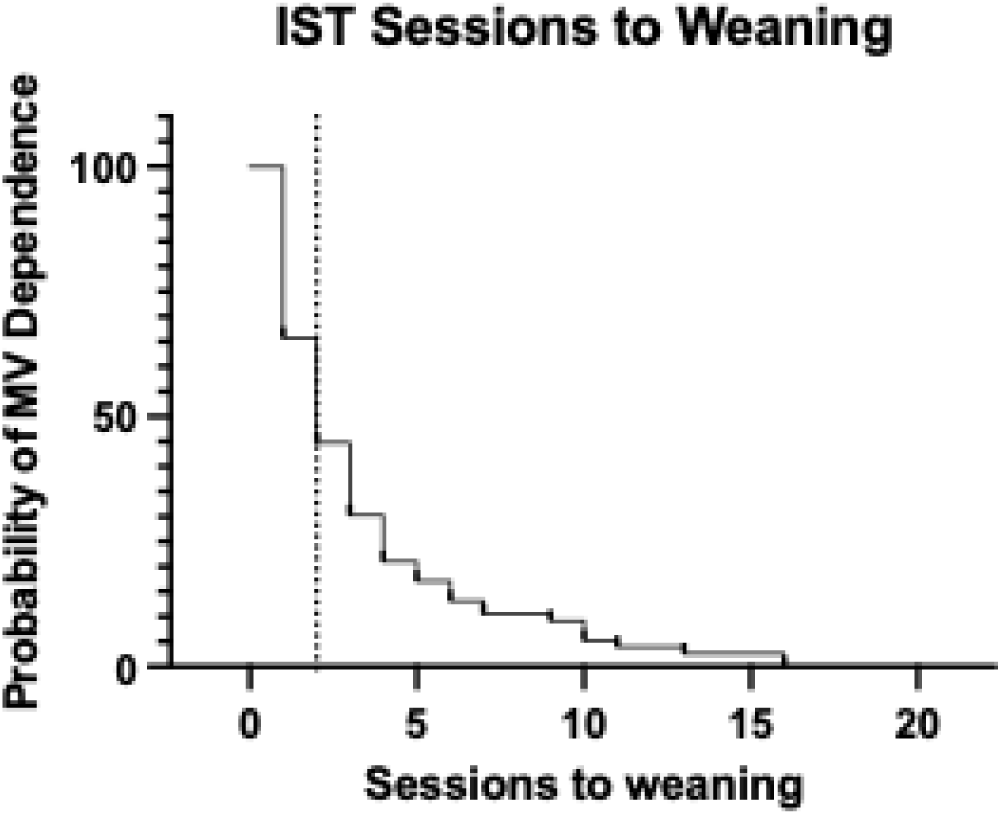
Analysis of Sessions to Weaning. Time-to-event curve illustrating successful weaning as a function of inspiratory strength training sessions. The median time to weaning was 2 sessions.

**Figure 4.**
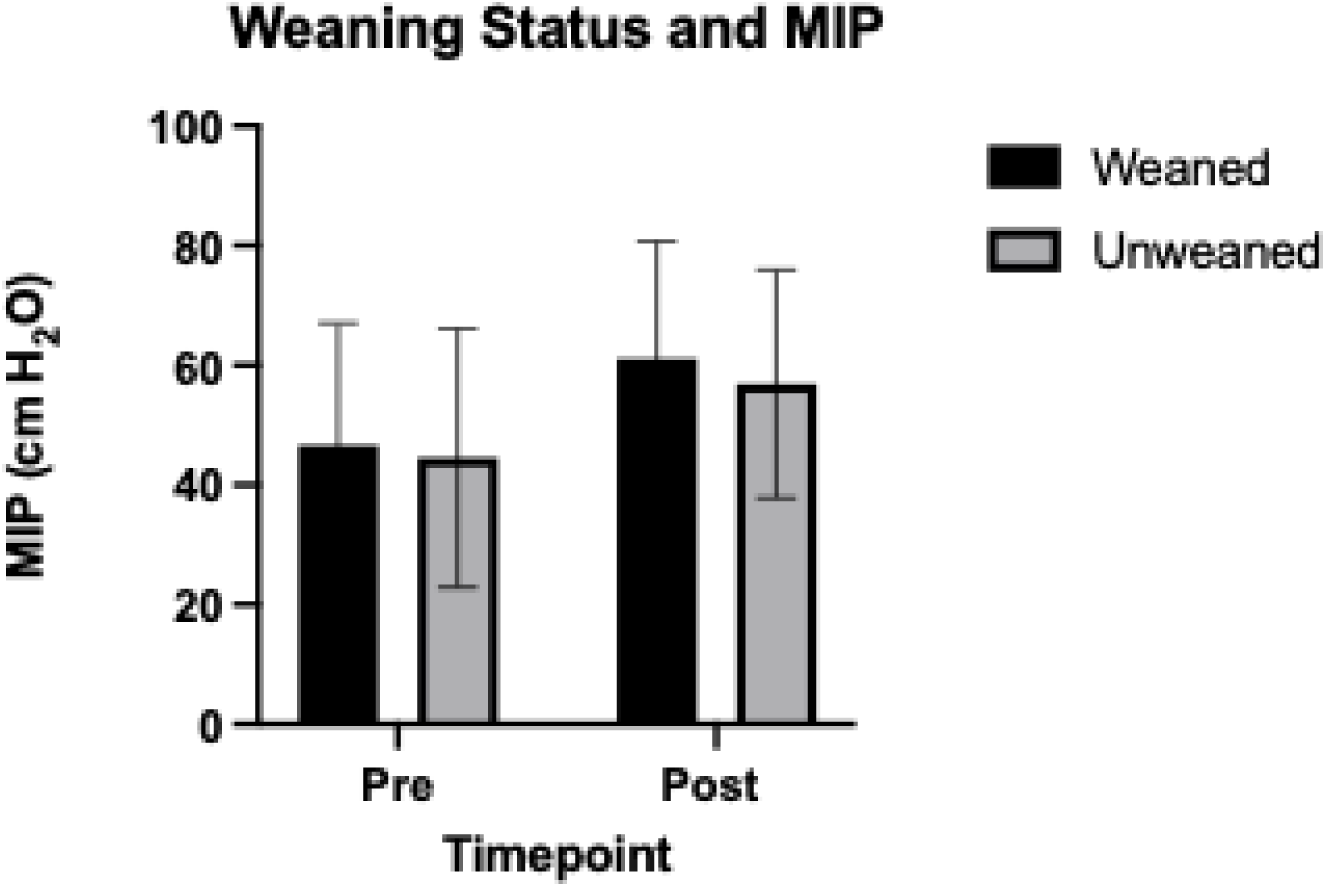
Comparison of Maximal Inspiratory Pressure According to Weaning Status. Mean maximal inspiratory pressure (MIP) values in weaned and unweaned patients. All patients demonstrated improvements regardless of weaning status.

### Evolving referral pattern over time

Case volume showed a non-significant upward trend, increasing by 2.49 case per year (R^2^ = 0.51, p=0.113, CI -0.93 to 5.9) (Figure 5). Median ventilator days prior to IST decreased from 23 (IQR 11.5-45.5) in 2015-2016 to 10 (IQR 6-26) in 2019-2021; however, this trend was not statistically significant (p=0.429). Similarly, median hospital days prior to IST declined from of 45 (IQR 21-59) in 2015-2016 to a median of 21 (IQR 9.5-48.5) (p=0.424). The PRISM IV scores (p=0.668), age at IST (p=0.589) and the S/F ratio (p=0.636) at the outset of IST remained unchanged across all time points. The number of IST sessions until weaning decreased significantly from a mean of 7 (SD 10.62) in 2015-2016 to 3.25 (SD 3.65) in 2019-2021 (p=0.011).

**Figure 5.**
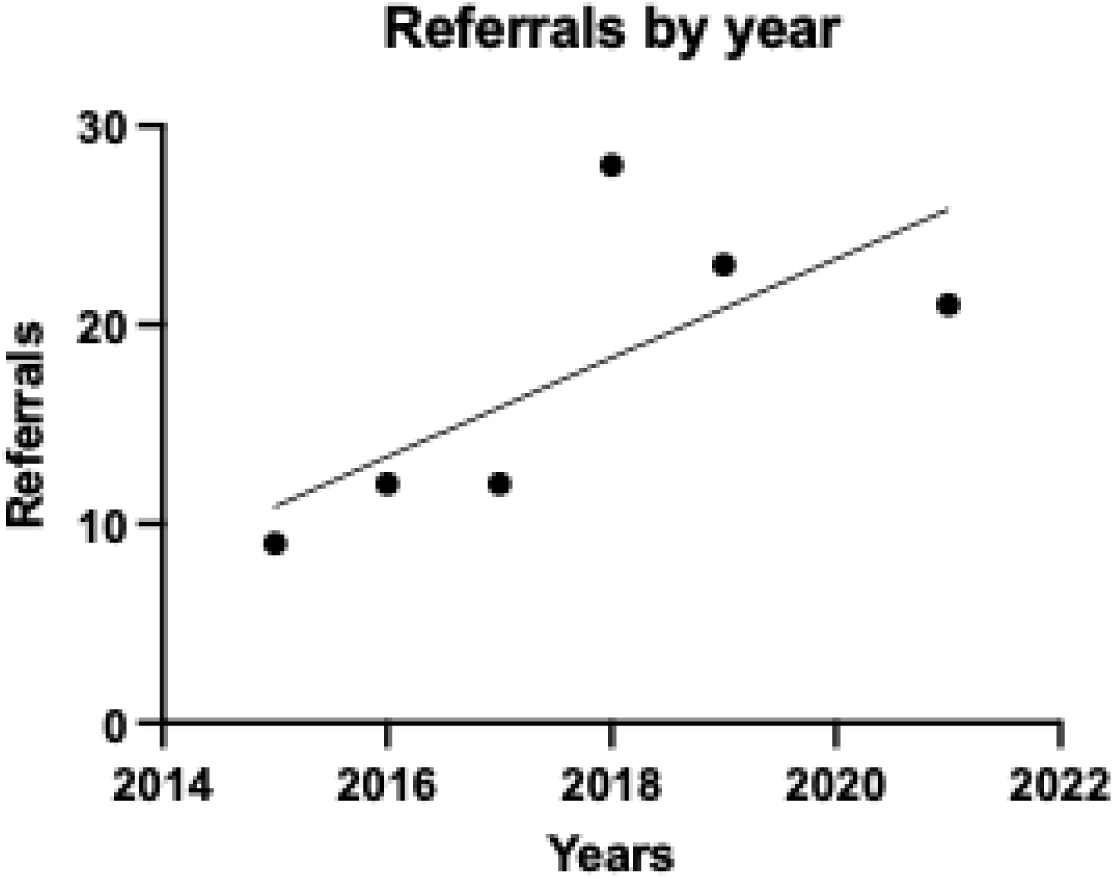
Annual Referral Volume over the Study Period. Number of patient referrals per year demonstrating changes in the referral pattern across the study period.

## Discussion

There is a vast body of evidence supporting the use of IST in the adult setting to facilitate ventilator weaning, and a small but growing body of evidence supporting its utility in the pediatric setting. To our knowledge, the only studies describing IST in the pediatric critical care setting to promote ventilator weaning are limited to small sample sizes or case reports. To date, this appears to be the largest reported dataset describing IST in the pediatric critical care setting. As advances in pediatric cardiac critical care continue to evolve, a growing number of children require intensive care and potentially prolonged bouts of mechanical ventilation. Prolonged mechanical ventilation, described as greater than or equal to 13 days in the pediatric literature, is tied to increased morbidity and mortality. IST is one potential strategy to preserve diaphragm motor function and respiratory drive, to enable more rapid ventilator weaning. Efficient ventilator weaning could be vital in improving outcomes for critically ill children.

In this single-center retrospective cohort, inspiratory strength training was feasible and well tolerated in a pediatric cardiac intensive care population with prolonged or complicated courses of care. IST was performed safely within the constraints of routine clinical practice by specially trained physical therapists who followed institutional practice guidelines. Despite a high illness acuity, nearly 90% of referred patients completed at least one session, and all monitoring events were transient with none required an escalation of care, supporting the feasibility of incorporating IST into pediatric cardiac ICU clinical practice. This is particularly important given the clinical complexity of this population. Among patients completing two or more sessions, maximal inspiratory pressure (MIP) increased from baseline to the final measurement. As the largest report to date of pediatric IST, it suggests IST is a clinically deliverable intervention that can produce measurable improvements in inspiratory muscle strength during critical illness.

Respiratory muscle weakness is increasingly recognized as a modifiable contributor to prolonged ventilator dependence in critically ill children. MIP has been used as a clinical indicator of inspiratory muscle strength and physiologic readiness for weaning in the pediatric critical care setting. There is substantial evidence in the adult literature supporting the use of IST to improve MIP in ventilated adults. Despite this, to date there remain limited reports of IST improving MIP in ventilated children. In this cohort, MIP improved in all groups receiving IST, regardless of weaning or survival status. Although spontaneous recovery, improvements in overall clinical status, and changes in sedation likely contributed to strength gains, the consistent improvement in MIP observed across patients supports the plausibility of IST as a targeted strategy to address respiratory muscle dysfunction in the pediatric cardiac ICU. The median of 2 IST sessions to achieve weaning is unlikely to have produced meaningful respiratory muscle hypertrophy or substantial improvements in contractile capacity. Instead, the applied inspiratory load may have augmented phrenic motor drive. During eupnic breathing, only approximately 30-50% of the phrenic motor pool is recruited.^29^ Mechanical ventilation has been shown to suppress phrenic motor output, thereby diminishing diaphragm activation.^30^ Prior work demonstrates that the application of an inspiratory load increases both burst frequency and duration of the phrenic motor neurons.^29^ This mechanism may underlie the observed improvements and warrents further investigation.

Liberation from invasive mechanical ventilation in the unique population appears multifactorial and not solely dependent on respiratory muscle strength. Patients who were not liberated from invasive mechanical ventilation had longer pre-intervention ventilator exposure and deeper sedation at IST initiation, suggesting greater illness severity and reduced readiness for weaning. Importantly, improvements in MIP were reported in both the weaned and unweaned groups, indicating that factors extending well beyond inspiratory weakness can contribute to weaning failure, such as ongoing renal or cardiac dysfunction, diffusion limitations…. (cites?). These findings highlight the importance of using IST as one component of a comprehensive multidisciplinary approach to ventilator weaning rather than as a stand-alone intervention.

The training technique utilized in our unit differs from the threshold technique reported in the majority of the adult literature and the vast majority of the existing pediatric literature. While some of our subjects performed training with a threshold device, 96% of patients trained received IST via an occlusive training approach. While threshold training devices are suitable for larger children, the physiologic dead space precludes the appropriate use of these devices in infants and small children with lower lung volumes. For this reason, the occlusive technique was the preferred training method in our study. The occlusive training approach was described and successfully utilized by Smith et al. in the Pediatric Cardiac ICU setting to promote weaning in a critically ill patient after cardiac surgery.^20^ This approach allows us to provide a strengthening intervention as well as document changes in MIP over time. Airway occlusion pressure has been shown to be an effective measure of patient effort and has been well correlated with esophageal pressure in children of all ages.^31^ While effective in eliciting maximal inspiratory effort, the Müeller maneuver has been associated with fluctuations in heart rate, blood pressure, arterial carbon dioxide and oxygen levels, and sympathetic drive.^32^ Given the potential for physiologic and hemodynamic changes, careful, continuous monitoring is required, along with extensive staff training and mentorship.

While our data demonstrated that IST is well tolerated, feasible, and yields a promising physiologic response when performed by a trained physical therapist, there are several limitations to our study design. This study is limited by its retrospective design and by the inclusion of only a specific subset of patients referred for IMT, which introduces selection bias and limits causal inference. Due to the nature of clinical documentation, data on missed sessions were unavailable. The cohort is heterogeneous with respect to diagnosis, age, illness severity, and sedation practices, and weaning decisions were not standardized. Future prospective studies should use standardized IMT protocols and prespecified outcomes (e.g., time to ventilator liberation, ICU/hospital length of stay, functional status at discharge). Despite these limitations, our study represents one of the largest descriptions of IST use in pediatric cardiac critical care and provides important real-world data supporting its feasibility and physiologic impact. While the body of literature on pediatric IST is growing, there remains a void in the literature for critically ill children. Further studies are needed to investigate the efficacy of IST in the PICU and PCICU settings and the utility of IST in facilitating more rapid ventilator weaning in critically ill children. These studies should relate results to overall hospital outcomes, such as length of stay, functional and feeding status upon discharge, and discharge disposition.

## Data Availability

Data Available upon request

## Acknowledgements

University of Florida Clinical and Translational Science Institute. This project was supported, in part, by the National Institutes of Health’s Clinical and Translational Science Institute under award number UL1TR001427. The content is solely the responsibility of the authors and does not necessarily represent the official views of the National Institutes of Health.

